# Understanding Social Ecological Factors of Firearm Safety Engagement Among Latino(a/e/x) and Hispanic Adults Near Albuquerque, New Mexico: a Concurrent Mixed-Methods Study

**DOI:** 10.64898/2026.03.24.26349234

**Authors:** Richardson Monte-Angel, Carmen Logie, Tanya Sharpe, Samantha Teixeira

## Abstract

**Background:** Disparities in injury and death indicate that Latinos and Hispanics are disproportionately affected by firearm violence. Understanding the factors that impact Latino and Hispanic engagement with firearm safety is integral to developing nuanced interventions, yet these factors remain largely understudied. This study explores the social ecological factors related to firearm safety engagement among Latino and Hispanic adults residing in New Mexico.

**Methods:** The study used a convergent mixed-methods design with quantitative and qualitative components. Data were collected from a predominantly Latino-Hispanic community experiencing high rates of firearm violence near Albuquerque, New Mexico. Quantitative data (n=303) were collected using a community-based survey with a non-random sample on firearm safety engagement, collective efficacy, and sociodemographic characteristics. Qualitative data (n=18) included semi-structured interviews from a subset of the survey population who expressed interest in participating. Quantitative data was used to explore descriptive statistics and correlations between reported levels of collective efficacy and firearm safety engagement. Qualitative data were used to explore the firearm safety experiences of Latino and Hispanic participants.

**Analyses:** Multivariate regression analyses examined associations between collective efficacy (exposure) and engagement with firearm safety (outcome). I also explored associations across key domains: collective efficacy, neighborhood characteristics, individual characteristics, and sociodemographic factors. Interviews were analyzed using framework analysis to generate a cohesive thematic structure informed by a social ecological model. The results from the quantitative and qualitative data were then integrated to develop a robust understanding of social ecological factors related to firearm safety engagement using a mixed methods joint display.

**Results:** There were 303 survey participants (40.6% male; 55.1% female; 4.3% other gender identity) and 18 interview participants in this study. 57.1% of survey participants reported engaging with at least one firearm safety practice or initiative. Results from multivariate regression indicated that higher collective efficacy (β = 0.082, p = 0.002), higher informal social control (β = 0.174, p = 0.001), stronger endorsement of gun safety principles (β = 0.079, p < 0.001), being married vs. unmarried (β = -0.334, p < 0.001), speaking Spanish in the home vs. English (β = 1.048, p < 0.001), and not owning a gun (β = - 0.638, p = 0.006) were significantly and positively associated with firearm safety engagement. Themes from the qualitative data included barriers (insecure environment; lack of meaningful engagement) and facilitators (location-specific contributors to safety; collective identity and pride) to firearm safety engagement, organized by social ecological domain. Mixed methods findings indicate factors associated with participants’ individual firearm safety engagement, while providing insights into the perceived barriers and facilitators across social ecological domains.

**Discussion:** Findings from this mixed-methods study suggest that processes of empowerment and collective efficacy may contribute to greater firearm safety engagement within Latino and Hispanic communities. Findings expand injury prevention research by exploring the factors influencing firearm safety engagement among a marginalized and hard-to-reach population who have disproportionate experiences with firearm victimization, perpetration, and injury.

**Conclusion:** This study offers unique methodological approaches by using concurrent mixed methods and collecting complementary data sources to understand firearm safety engagement among Latinos and Hispanics. Findings highlight the need for culturally specific and community-engaged interventions that address social ecological disparities to strengthen safety practices and reduce firearm-related harms.

## 1. Introduction

Of all fifty states, New Mexico has consistently ranked highest for gun deaths and suicide among Latino and Hispanic people (1). In 2022, 63% of New Mexican firearm homicide victims were Latino or Hispanic (2). In comparison, non-Hispanic white residents of New Mexico made up only 21% of firearm homicide victims. This is concerning given that nearly half of New Mexicans identify as Latino and/or Hispanic at 49.2% (3). In response, calls have been made for approaches to increase engagement with firearm safety, including education on safe usage, storage, and gun safety policies (4).

Firearm safety can be better understood by examining social ecological factors, described as the broad range of elements within social structures and ecological systems that influence behaviors, health, or outcomes (5). Social ecological factors have been previously analyzed across interconnected societal domains to better understand firearm violence (6, 7). However, seldom has research investigated the social ecological factors that impact engagement with firearm safety.

To address this gap in the literature, this study employs a concurrent mixed methods approach to examine the social ecological factors impacting firearm safety engagement among Latino and Hispanic adults residing in New Mexico. To achieve this objective, this study adopts a social ecological framework conceptualizing firearm safety engagement as being influenced by intrapersonal, interpersonal, cultural, communal, organizational, and structural domains.

### 1.1 Social ecological factors of Latino-Hispanic firearm safety engagement

#### 1.1.1 Intrapersonal factors

There are multi-level social ecological factors that may impact firearm safety engagement. In the intrapersonal domain, prior research suggests that firearm safety engagement may be associated with underlying beliefs, perceived threats to safety, and gun ownership (8, 9, 10,11). Underlying beliefs including trust in the government and a need for protection have been previously associated with distinct firearm behaviors and safety practices (12, 13, 14). For instance, a cross-sectional study using a nationally representative sample of US adults (n=10,000) to examine factors related to secure firearm storage found that individuals who owned firearms primarily for protection were less likely to engage in secure storage than those who owned them for hunting or sport (12). However, empirical studies about the underlying attitudes and beliefs associated with firearm safety engagement are limited in terms of publications, and calls have been made to address this gap in the literature (12, 9, 14).

Perceiving threats to safety has also been previously associated with distinct firearm practices (15, 16). For instance, a cross-sectional study study using data from a nationwide survey found that gun owners who experienced higher levels of sociocultural anxiety were more likely to always have a loaded firearm accessible at home, store firearms locked and loaded, and believe that firearms make homes safer (11). Perceived neighborhood disorder, defined as residents’ interpretations of signs in their communities that suggest a breakdown in social control, has also been associated with distinct firearm behaviors (17, 18, 19, 20). For example, one recent study using longitudinal data of adolescents (n=11,887) found perceived neighborhood disorder to be associated with a higher prevalence of gun carrying (18). Further, beliefs about firearm safety have been shown to differ based on previous gun violence experiences, particularly among marginalized groups (10, 17). For instance, one recent cross-sectional study surveyed a multiracial sample of youth (n=276) living in New Orleans, Louisiana to examine the association between youth gun beliefs and previous gun violence exposure (10). Among Latino-Hispanic youth in this study, indirect gun violence exposure was strongly associated with positive beliefs about gun ownership and public carriage (OR = 3.95, 95% CI [1.01, 15.50]). While this and other studies have investigated the underlying beliefs associated with firearm practices, not often have these beliefs been examined in relation to firearm safety engagement among Latinos and Hispanics.

Support for firearm safety practices and initiatives may also be distinct for Latino and Hispanic gun owners compared to other racial and ethnic groups. For example, a cross-sectional study using data from a nationally representative sample of US adults examining public support for firearm policies (n=2,778) found that Hispanic gun owners were more likely than other racial groups to support legal carrying, but less likely to believe that owning a gun improved their personal safety (21). Recent qualitative research also indicates that minority gun owners may perceive the need to form their own social communities based on a desire for self-protection from other citizens and law enforcement (22). The firearm safety beliefs of Latinos and Hispanics are particularly important to understand as gun ownership in these communities is on the rise. In 2020, one-fifth of new US gun owners identified as Latino (23). Knowledge of how gun ownership influences firearm safety engagement is also essential due to the heightened risk of experiencing firearm-related harms for both Latinos and Hispanics (2) and gun owners (12, 21). However, few studies have examined how owning a gun may impact Latino and Hispanic firearm safety engagement.

#### 1.1.2 Interpersonal and cultural factors

Within the interpersonal and cultural domains, family structure and cohesion, defined as positive relationships and connectedness within the family unit, have been previously associated with differences in firearm safety behaviors and beliefs (24). Recent studies have found that family cohesion may mitigate unsafe firearm behaviors, and some Latino and Hispanic subgroups have been considered to be more family-oriented compared to whites due to being more focused on family rather than individual well-being (25, 26, 27, 28). For instance, one cross-sectional study using a nationally representative sample to identify risk and protective factors for weapon involvement among African American, Latino, and white adolescents (n=13,503) found family connectedness to be protective against weapon involvement only for Latino adolescents (28).

Marital status is another aspect of family structure that has been previously shown to influence differences in firearm-related behaviors, including increased household firearm ownership (29, 30). This relationship has been shown to overlap with other demographic variables, including gender and the presence of children in the home. One cross-sectional study examining the parental motivations for keeping firearms in the home (n=2,924) found that households with married parents/caregivers were at an increased likelihood for owning firearms compared to younger parents/caregivers with less education [OR = 1.29, p < 0.05] (30). Nuanced research involving the family structures and marital status of Latino and Hispanic individuals may provide deeper insight into the factors associated with firearm safety engagement.

#### 1.1.3 Communal factors

Within the communal domain, racially homogenous neighborhoods, where the majority of residents belong to the same racial or ethnic group, may also affect the firearm experiences of Latinos and Hispanics (31, 32). Social disorganization theory posits that racially homogeneous neighborhoods experience decreased levels of violence, crime, and neighborhood disorder due to increased social cohesion (31, 33, 34). For instance, a cross-sectional study using data from 2,720 households with low- to moderate-income families found that respondents in racially homogeneous neighborhoods exhibited higher degrees of social cohesion and collective efficacy (35). These benefits have been most often observed in areas of high immigrant concentration, where a mutual understanding of the challenges faced in learning a new language and adopting customs of the host country may deepen community commitments (33, 36). This finding may be particularly salient for Latino and Hispanic communities, as US immigrants predominantly migrate from Latin-American countries, particularly Mexico and Cuba (37). However, little research has been conducted exploring Latino-Hispanic racial homogeneity in relation to firearm safety engagement.

Some scholars have questioned the role that racial neighborhood homogeneity plays in protecting communities against violence (35, 38), arguing that focusing solely on racial homogeneity detracts from potentially confounding factors that may influence cohesion within a community, such as immigration status and concentration, concentrated neighborhood disadvantage, or the availability of communal resources (39, 40). Previous research also suggests that in some contexts, neighborhood racial homogeneity may increase the likelihood of Latinos and Hispanics experiencing other forms of violence, such as police violence (38, 41). The rapid growth of Latino-Hispanic populations in the US, coupled with disproportionately high rates of firearm violence and neighborhood disadvantage experienced by these groups, merits further investigation of the impact of racial neighborhood homogeneity on firearm safety (42). However, this connection has yet to be explored.

#### 1.1.4 Organizational factors

The inability to comprehend firearm safety resources may be a significant organizational barrier to firearm safety engagement among marginalized groups, including Latinos and Hispanics (43, 44). Health literacy, defined as the ability to find, understand, and use information and services to inform health-related decisions, is impacted by the accessibility and clarity of health and safety resources (45). US Latinos and Hispanics who cannot speak English proficiently may face healthcare obstacles because of the predominant use of English in healthcare and lack of Spanish-language resources (46, 47). Recent studies indicate that health literacy challenges may be ubiquitous among firearm safety resources, resulting in recurring firearm violence experiences for individuals who cannot comprehend them (46, 48). For example, a recent study analyzing the comprehensibility of firearm-safety resources at verified trauma centers, national health organizations, and gun violence prevention advocacy groups found that only 21% of hospitals and 22% of national online injury prevention resources related to firearm safety met the sixth-grade reading level recommended by the American Medical Association (46). This disparity is particularly concerning for the more than 28% of US Latinos and Hispanics who reported speaking and comprehending English less than “very well” (49). However, not often has research investigated the impact that Spanish language use may have on firearm safety engagement.

#### 1.1.5 Structural Factors

Structural inequities such as concentrated disadvantage and infrastructure challenges may create barriers to firearm safety engagement within Latino and Hispanic communities. Social disorganization theory suggests that neighborhoods with insufficient economic resources, poor residential participation in community efforts, and deficient infrastructure provide opportunities for firearm violence (50). High rates of violence and poor health outcomes then concentrate disadvantage, perpetuating cycles of violence (51). This is a significant issue for Latinos and Hispanics in the US, who disproportionately reside in socioeconomically disadvantaged communities (52, 53). In 2021, 18% of US Hispanics were living in poverty, almost three times higher than non-Hispanic whites (54). While disparities in firearm violence experiences for Latinos and Hispanics residing in disadvantaged communities have been studied (55, 56), little research has been conducted that examines the potential impact of concentrated disadvantage on firearm safety engagement.

### 1.2 Facilitators of firearm safety engagement: empowerment and collective efficacy

Empowerment and collective efficacy are community-level factors which may facilitate greater firearm safety engagement (57, 58). Empowerment, defined as a process situated within social inequity that aims to shift power relations, has been shown to occur at both individual and community levels (59). Individual empowerment is concerned with the ability of a person to incite collective action (60). Collective empowerment is concerned with communal factors that can empower individuals and groups to act (61). Empowerment-based interventions have found success in engaging diverse groups with firearm safety (62, 63, 64, 65). One such intervention was developed to empower loved ones of individuals at risk for suicide (n=23) to discuss secure firearm storage for suicide prevention with their loved ones (62). This intervention resulted in a 90-minute workshop with participants reporting enhanced willingness to discuss storage with their loved ones (62).

While empowerment has been associated with increased firearm safety engagement, it has also been found to engender a greater sense of safety and confidence among gun owners, resulting in unique shared identities among some groups (22, 66, 67). For example, a recent study using data from an original Mechanical Turk survey (n = 876) examined the extent to which gun owners found guns to be emotionally and morally empowering (68). The results of this study indicated that gun owners viewed guns themselves as emotionally and morally empowering irrespective of their fears of crime, prior victimization, police dissatisfaction, and political attitudes (68). Empowerment may similarly play a complex role in the firearm safety practices of diverse groups, although this remains empirically unexplored.

Collective efficacy, defined as a community’s shared belief in its ability to take actions to produce change, has been previously associated with reductions in firearm violence (69, 70, 71, 72). A recent cross-sectional study examining the association of collective efficacy and exposure to firearm homicide among adolescents (n=1,736) found that exposure risk decreased with greater neighborhood collective efficacy (69). Conceptually, collective efficacy is often divided into two constructs: social cohesion (SC), referring to the norms, values, and mutual trust among neighbors, and informal social control (ISC), reflecting residents’ willingness to intervene to prevent harmful behaviors (72). Previous research has shown collective efficacy to be associated with increased behaviors that work to improve communities, including civic engagement (70, 71), volunteering (73), mobilization against crime (74), and bystander intervention (75). Collective efficacy may also be associated with greater firearm safety engagement, although this relationship has yet to be empirically investigated.

### 1.3 Current study

This study adopts a concurrent mixed methods design to address a critical gap in the literature by exploring the social ecological factors of firearm safety engagement among Latinos and Hispanics in New Mexico. The mixed methods design is anticipated to be useful in providing both depth and breadth of this underexplored topic area with participants originating from an understudied community. The mixed methods research question was: ‘How do Latino and Hispanic adults residing in a community near Albuquerque, New Mexico perceive firearm safety engagement?’ For the quantitative portion, it was hypothesized that collective efficacy and its subscales would be significantly and positively associated with increased engagement with firearm safety, with this association influenced by sociodemographic characteristics.

## 2. Methods

### 2.1 Study design

This study employed a concurrent mixed methods design (76). This design begins with quantitative data collection and analysis, followed by qualitative data collection and analysis to complement and expand the findings of the quantitative results (77). Specifically, in the first phase of the study, cross-sectional survey data (n=303) was collected and analyzed to investigate the associations between firearm safety engagement, sociodemographic characteristics, and collective efficacy. The second phase was qualitative inquiry with survey participants who expressed interest and volunteered their contact information (n=18) to provide contextual information on the barriers and facilitators to firearm safety engagement. Participants were eligible for this study if they (1) identified as Latino(a/e/x) and/or Hispanic; (2) were 18 years or older; and, (3) identified as residents of the target community. Both written (survey) and verbal (interviews) informed consent was obtained from participants prior to survey and interview completion.

Following concurrent mixed methods design, I separately analyzed quantitative and qualitative data and then integrated data strands together (76). Methodological integration occurred through merging and connecting the two strands of data. Merging was accomplished during the study design phase, when interview questions were developed to contextualize the scales and instruments included on the survey. Connecting was achieved by recruiting interviewees from the sampling frame of survey participants. The scales on the survey assessed firearm safety engagement as it relates to perceived collective efficacy, neighborhood disorder, underlying beliefs regarding firearms, and sociodemographic characteristics. Interview questions were developed to elicit how participants perceived their community’s engagement with firearm safety initiatives and practices. Figure 1, ‘Procedural Diagram for Concurrent Design Procedures’, outlines the procedural steps of developing, modifying, and integrating data in the phases of the study.

**Figure 1.**
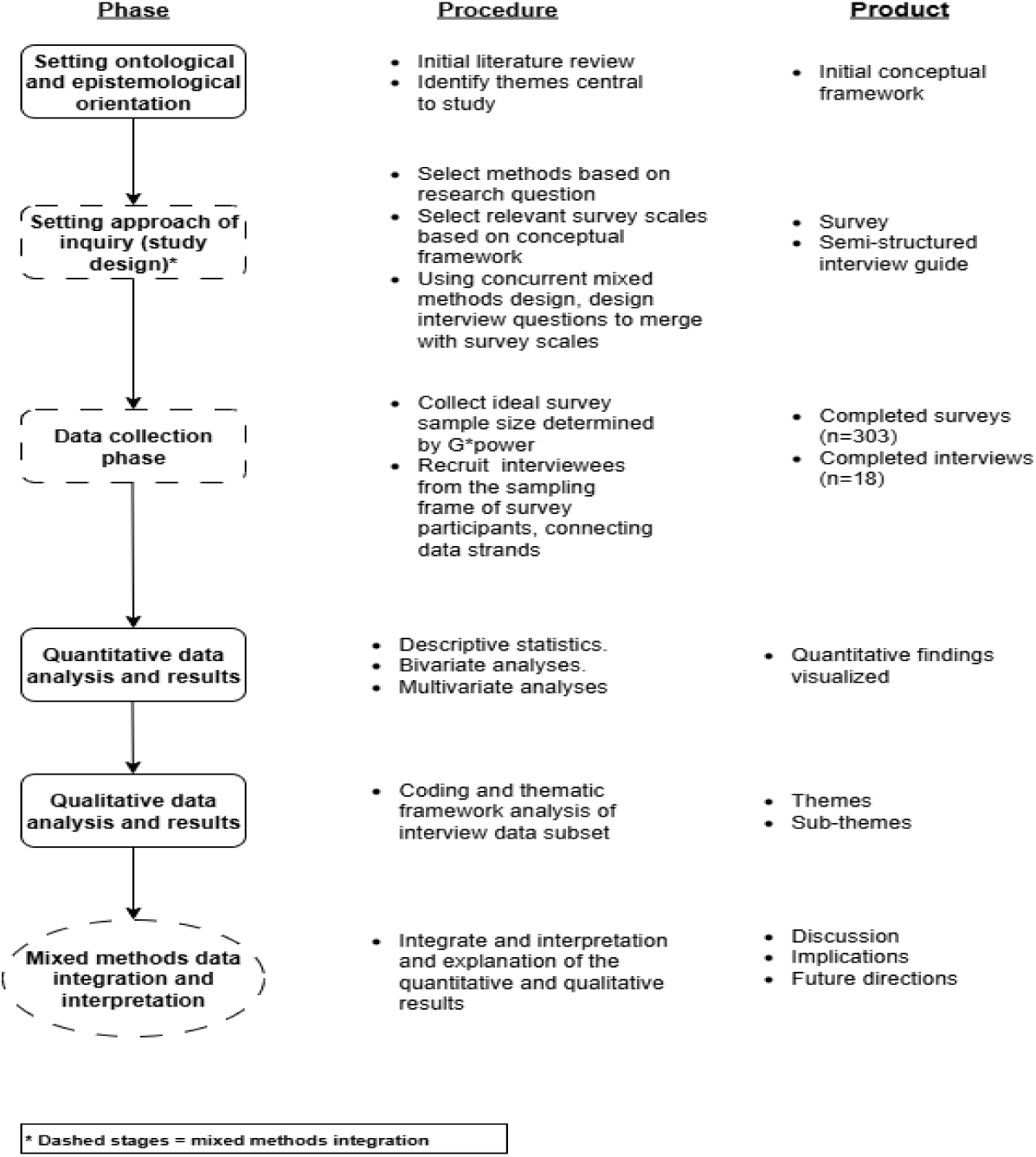
Procedural Diagram for Concurrent Design Procedures

### 2.2 Setting

I collected all data in a community near Albuquerque, New Mexico from April to September of 2024. I selected this community as the study site for several reasons. First, firearm violence has been shown to substantially impact this community and is widely reported by the media. Second, firearm safety and prevention efforts are often targeted towards this community. Third, Latinos and Hispanics comprise 81% of the community’s residents, indicating relatively high ethnic and racial homogeneity (78).

### 2.3 Data collection

I recruited survey participants using non-random snowball sampling methods (79), including distributing flyers throughout the target community, administering surveys in-person, and posting study announcements on social media platforms, especially those connected to local community initiatives. Before launching data collection, I built relationships with community-based organizations to build trust and secure collaboration, including public health outreach groups and Spanish-speaking coalitions. I administered the survey both online and in-person, offering it in English and Spanish to accommodate participant language preferences. Upon survey completion, participants received a $10 gift card, either physically (for in-person participation) or electronically (for online participation).

I augmented the sample using respondent driven sampling (RDS), a method of data collection involving participants who initially completed the survey and agreed to receive small compensation to recruit others who met inclusion criteria (80). For this study, twelve participants agreed to serve in this role, and were given the opportunity to receive up to ten coupons worth $10 each to recruit participants.

The ideal survey sample size was determined to be equal to or greater than 300, determined using G*power analysis, taking the expected total Latino-Hispanic population over 18 in the target community and calculating for a 95% confidence interval and 5% margin of error (81). Less than half (n=120, 39.6%) of participants completed the survey using personal links provided to study ‘seeds’ using RDS. A small number of participants (n=36, 11.8%) completed paper surveys via in-person recruitment, which were then entered manually. The remaining participants (n=147, 48.5%) completed the survey using an anonymous link or QR code. As two forms of non-random sampling were used, weighting was not applied.

Following completion of the survey, participants were invited to participate in one-on-one interviews. In total, 18 individuals participated in interviews from June to September of 2024. The interviews lasted up to 45 minutes and were conducted in either English or Spanish, in-person in a private space or via a web video platform at the preference of participants. Participants were informed the interview would be recorded and assured that no identifying information would be collected. All participants enrolled following verbal and written informed consent, and each was paid a $20 gift card as compensation.

### 2.4 Quantitative measures

The quantitative measures in this study included the outcome variables (1) awareness of, and (2) involvement in firearm safety practices and initiatives, and exposure variables including collective efficacy, neighborhood characteristics, individual characteristics, and sociodemographic factors.

#### 2.4.1 Outcome variables

The outcome variables measuring firearm safety engagement were (1) gun safety involvement, and (2) gun safety awareness, operationalized by an index of binary outcomes and Likert scale, respectively, created for the present study. Both the index and scale assess engagement with firearm safety practices and initiatives across five domains: (1) safe and secure gun storage; (2) reducing firearm availability for children and people at risk of harming themselves and others; (3) holding the gun industry accountable to ensure adequate oversight; (4) engaging responsible gun dealers and owners in solutions, and; (5) insisting on mandatory training and licensing for gun owners (82, 83, 84). Items were identified from previous firearm safety resources and research pertaining to firearm safety screening, and included items such as, “Have you ever discussed with children what to do if they find a gun?” (21, 85, 86). Participants were asked about their involvement (‘Yes’, ‘No’, ‘Not Applicable to me’) and awareness of these practices and initiatives (‘Very unaware’, ‘Unaware’, ‘Aware’, ‘Very aware’) separately. Empirically separating the potential for firearm safety engagement (awareness) and actual exercise of firearm safety (involvement) permits a more nuanced examination that accounts for potential associations with collective efficacy (87). Internal consistency was acceptable for both scales (Involvement: α = 0.788; Awareness: α = 0.844).

#### 2.4.2 Exposure variables

The exposure variables included collective efficacy, neighborhood characteristics, individual characteristics, and sociodemographic factors. Collective efficacy was operationalized using the social cohesion and informal social control scales developed by Sampson et al (72), applied in recent research measuring collective efficacy in Latino and Hispanic populations (88). Social cohesion was measured using four items, including, “People in this neighborhood can be trusted.” Factor loadings from the measurement model have been shown in previous research to range from 0.832 to 0.963. For the current study, acceptable internal consistency was also reached (α = 0.828). Informal social control is measured using four items to assess residents’ perceptions of the likelihood that neighbors would intervene if they observed negative situations such as: “If a child was disrespecting an adult.” Loadings from the measure have been shown to range from 0.717 to 0.995. For the current study, acceptable internal consistency was also reached (0.762).

Neighborhood characteristics examined as exposures included Perceived Neighborhood Disorder (PND). This was measured with the Perceived Neighborhood Disorder Scale (PND, 19), a 15-item self-report measure of physical and social order in a community applied in recent research in diverse populations (89). Sample items include, “There is too much drug use in my neighborhood.” Scores are summarized into a total score (range 15–60), and the scale has been found to be a reliable and valid measure with factor loadings ranging from 0.893 to 1.035. For the current study, acceptable internal consistency was reached (0.776).

Individual characteristics examined as exposures included principles of firearm safety and gun ownership status. Developed by Grene et al (9), the ‘Principles Underlying Opinions on Gun Safety’ scale assesses the principles that people hold regarding firearms with a 5-point Likert scale. Items include, “People who have been convicted of a violent crime should not be able to purchase or possess a gun”, and acceptable internal consistency was reached for the current study (0.807). This scale has been previously employed by Grene et al (9) to assess gun owners’ perceptions of firearm safety policies (α = 0.861). Gun ownership status was assessed with a binary question, “do you or does anyone in your household own a gun?” Due to known potential legal implications of marginalized groups possessing firearms, participants were able to opt out of answering this question by answering, ‘prefer not to say’ (n=15, 4.9%).

Lastly, sociodemographic factors examined as exposures included age, gender, ethnicity, country of origin, marital status, highest level of educational attainment, gross household income, housing status, and language spoken in the home.

## 3. Analytical Approach

### 3.1 Statistical methods

Descriptive statistics and chi-square (χ²) tests were used to compare variables across key domains: collective efficacy, neighborhood characteristics, individual characteristics, and sociodemographic factors. Variables showing statistically significant associations (p < 0.05) at the bivariate level were included in multivariate stepwise regression models. Backward stepwise linear regression was used. Variables not demonstrating significant bivariate associations were excluded, consistent with recommended procedures for exploratory regression modeling (90). Unstandardized coefficients (β) and standardized coefficients (aβ) were reported, and all statistical analyses were performed using SPSS Version 29.

I conducted PCA and varimax rotation to assess construct validity. From the scale, two distinct components out of eight items were identified with eigenvalues greater than one, accounting for 65% of the variance. The first component included all items related to keeping guns out of the hands of high-risk groups and greater concern for the ubiquity of different forms of firearm violence. The second component included all items related to firearm ownership being a fundamental, unchangeable right, coupled with lower concern for the various forms of firearm violence. All items loaded >1.5 on one of the two components without cross-loading above 0.8. The resulting factors were labeled as: (1) Endorsement of Gun Safety to Reduce Gun Violence (GV) (α = 0.888); and, (2) Endorsement of Firearm Ownership as a Fundamental Right (α = 0.828).

### 3.2 Qualitative analysis

Using a semi-structured interview guide (91), participants were asked questions about their perceptions of the potential barriers to firearm safety engagement for themselves and their community. Aligning with the theoretical integration of this study, questions focused on factors across six social ecological domains. Questions were also asked to explore factors that may facilitate greater firearm safety engagement. Following interviews, I conducted personal reflections via journaling, which involved examining my impact throughout the study and noting the reactions of others to me (91). All interviews were recorded, de-identified and imported into NVivo v14. Transcripts were produced using Zoom transcription software and then corrected for accuracy using triangulation, involving multiple sources of data to cross-check findings and identify inconsistencies (92).

Transcripts were analyzed using framework analysis, a systematic qualitative method that proceeds through five interconnected stages to generate a cohesive thematic structure (93). The first phase, familiarization, involved immersing myself in the data through repeated reading of the 18 transcripts to gain an overall sense of key ideas and emerging patterns. In the second phase, I identified recurring codes that provided the foundation for an initial coding framework. In the third phase, I systematically applied this framework to the dataset by linking codes to specific quotes to create subthemes. In the fourth phase, I charted data to organize it into matrices, incorporating all themes, subthemes, codes, and illustrative quotes. In the final phase, I mapped and synthesized the charted data to explore relationships, refine categories, and generate insights, resulting in a rigorous, transparent framework. Themes were mapped by (1) social ecological domain, and (2) whether they presented as a barrier to or facilitator of firearm safety engagement.

## 4. Quantitative Results

### 4.1 Main results: multivariate analyses

See Table 1 for a full list of descriptive results. All variables found to be significantly associated to the outcome variables through Pearson’s correlation were included in subsequent multivariate analyses (see Table 2).

**Table 1.** Demographic characteristics of Latino-Hispanic survey participants in New Mexico.

| Characteristic | n | Valid % |
| --- | --- | --- |
| Gender identity |  |  |
| Cisgender woman | 167 | 55.1% |
| Cisgender man | 124 | 40.6% |
| Non-binary | 6 | 2% |
| Age (mean: 47.5 years; S.D: 15.7 years) |  |  |
| 18-24 | 38 | 12.5% |
| 25-34 | 78 | 25.7% |
| 35-44 | 75 | 24.7% |
| 45-54 | 34 | 11.2% |
| 55-64 | 33 | 10.8% |
| 65-74 | 32 | 10.6% |
| 75+ | 13 | 4.2% |
| Ethnicity |  |  |
| Hispanic | 106 | 35% |
| Mexican | 75 | 24.8% |
| Latine(a/o/x) | 56 | 18.5% |
| Mex-American | 29 | 9.6% |
| Other | 37 | 12.3% |
| Language of survey |  |  |
| English | 211 | 69.6% |
| Spanish | 92 | 30.4% |
| Gun ownership status |  |  |
| Yes | 128 | 42.6% |
| No | 157 | 52.5% |
| Prefer not to say | 15 | 5% |
| Country of origin |  |  |
| US | 169 | 55.8% |
| Mexico | 104 | 34.3% |
| Other | 30 | 9.9% |
| Language spoken at home |  |  |
| English | 145 | 47.9% |
| Spanish | 138 | 45.5% |
| English and Spanish | 20 | 6.6% |
| Housing status |  |  |
| Homeowner | 193 | 63.7% |
| Renter | 95 | 31.4% |
| Houseless | 15 | 5% |
| Educational attainment |  |  |
| Elementary school | 11 | 3.6% |
| Middle school | 11 | 3.6% |
| High school | 117 | 38.6% |
| Associate's/cert | 75 | 24.8% |
| Bachelor's degree | 66 | 21.8% |
| Graduate/doctoral degree | 22 | 7.3% |
| Income |  |  |
| < 20,000 | 68 | 22.4% |
| \$20,000 to \$40,000 | 89 | 29.4% |
| \$40,000 to \$60,000 | 74 | 24.4% |
| \$60,000 to \$80,000 | 44 | 14.5% |
| > \$80,000 | 28 | 9.2% |
| Marital status |  |  |
| Married | 143 | 47.2% |
| Widowed | 21 | 6.9% |
| Divorced | 29 | 9.6% |
| Partnered | 34 | 11.2% |
| Single, never married | 76 | 25.1% |
| Length of residency in community |  |  |
| < 6 mo | 4 | 1.3% |
| 6 mo to 12 mo | 7 | 2.3% |
| 12 mo to 2 yrs | 15 | 5% |
| 2 to 5 yrs | 34 | 11.2% |
| 10 to 20 yrs | 83 | 27.4% |
| > 20 yrs | 123 | 40.6% |

**Table 2.**
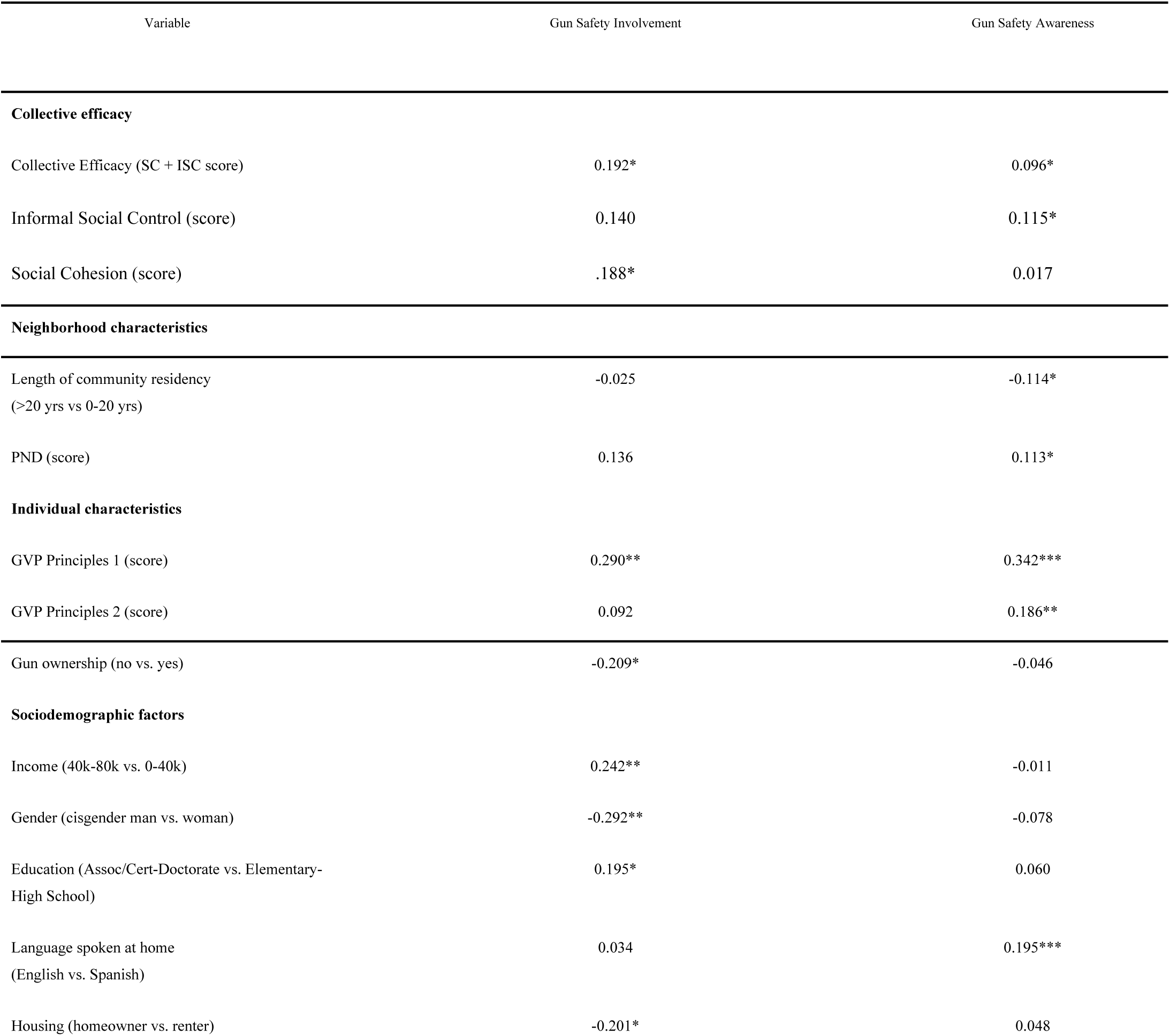
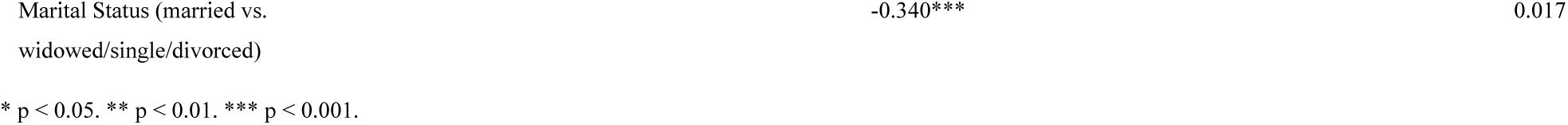
Pearson’s Correlations: Gun safety involvement & awareness among 303 Latino-Hispanic adults in New Mexico.

#### 4.1.1 Gun safety involvement

Variables associated with gun safety involvement at the bivariate level were entered into a backward stepwise linear regression model. Of these, four predictors were retained in the final model: (1) collective efficacy; (2) endorsement of gun safety to reduce gun violence; (3) marital status, and; (4) gun ownership status. The overall model explained 31.2% of the variance in gun safety involvement (R² = 0.312, adjusted R² = 0.287). The final stepwise iteration did not result in a statistically significant improvement in model fit (ΔR² = −0.016, F change(1, 111) = 2.706, p = 0.103).

Consistent with the study hypothesis, higher collective efficacy was associated with higher gun safety involvement (β = 0.082, 95% CI [0.030, 0.135], aβ = 0.252, p < 0.01). Endorsing gun safety to reduce gun violence was also significantly associated with higher gun safety involvement (β = 0.079, 95% CI [0.044, 0.114], aβ = 0.355, p < 0.001), as was being married relative to unmarried respondents (β = -0.334, 95% CI [−0.489, −0.179], aβ = -0.336, p < 0.001). Lastly, not owning a firearm was associated with greater involvement relative to firearm ownership (β = −0.638, 95% CI [−1.088, -0.189], aβ = -0.221, p < 0.01; see Table 3).

**Table 3.** Stepwise Multivariate Regression Predicting Gun Safety Involvement and Gun Safety Awareness.

| Model 1: Gun Safety Involvement | Unadjusted $\beta$ (SE) | 95% CI | Adjusted $\beta$ | p |
| --- | --- | --- | --- | --- |
| *Collective efficacy* |  |  |  |  |
| Collective Efficacy (SC + ISC score) | 0.082 (0.026) | [0.030, 0.135] | 0.252 | 0.002** |
| *Individual characteristics* |  |  |  |  |
| Gun Safety to Reduce GV (score) | 0.079 (0.018) | [0.044, 0.114] | 0.355 | < 0.001*** |
| Gun Ownership (no) | -0.638 (0.227) | [-1.088, -0.189] | -0.221 | 0.006** |
| *Sociodemographic factors* |  |  |  |  |
| Marital Status (married) | -0.334 (0.078) | [-0.489, -0.179] | -0.336 | < 0.001*** |
| Gender (cisgender man) | -0.197 (0.120) | [-0.435, 0.040] | -0.103 | 0.103 |
| Income (40k-60k) | 0.100 (0.115) | [-0.127, 0.328] | 0.076 | 0.383 |

| Housing (homeowner) | 0.143 (0.321) | [-0.493, 0.779] | 0.040 | 0.656 |
| --- | --- | --- | --- | --- |
| Education (Assoc/Cert-Doctorate) | -0.068 (0.128) | [-0.321, 0.184] | -0.054 | 0.593 |
| $R^2 = 0.318$; Adjusted $R^2 = 0.287$; $\Delta R^2 = -0.016$; $F \text{ change}(1, 111) = 2.706$, $p = 0.103$ | | | | |
| * $p < 0.05$. ** $p < 0.01$. *** $p < 0.001$. | | | | |
| Model 2: Gun Safety Awareness | Unadjusted $\beta$ | 95% CI | Adjusted $\beta$ | p |
| *Collective efficacy* |  |  |  |  |
| Informal Social Control (score) | 0.174 (0.054) | [0.067, 0.281] | 0.171 | 0.001** |
| Collective Efficacy (SC + ISC Score) | 0.025 (0.082) | [-0.135, 0.186] | 0.034 | 0.756 |
| *Neighborhood characteristics* |  |  |  |  |
| PND (score) | 0.029 (0.027) | [-0.025, 0.083] | 0.058 | 0.283 |
| Length of communal residency (> 20 yrs) | -0.201 (0.132) | [-0.462, 0.059] | -0.081 | 0.129 |
| *Individual characteristics* |  |  |  |  |
| Gun safety to reduce GV (score) | 0.189 (0.034) | [0.121, 0.256] | 0.314 | < 0.001*** |
| Gun ownership as fundamental right (score) | 0.105(0.057) | [-0.007, 0.217] | 0.105 | 0.065 |
| *Sociodemographic factors* |  |  |  |  |
| Language spoken at home (Spanish) | 1.048 (0.314) | [0.430, 1.666] | 0.179 | < 0.001*** |
| $R^2 = 0.182$; Adjusted $R^2 = 0.171$; $\Delta R^2 = -0.006$; $F \text{ change}(1, 295) = 2.316$; $p = 0.129$ | | | | |
| * $p < 0.05$. ** $p < 0.01$. *** $p < 0.001$. | | | | |

#### 4.1.2 Gun safety awareness

Variables associated with gun safety awareness at the bivariate level were also entered into a backward stepwise linear regression model. Of these, three predictors were retained in the final model: (1) informal social control; (2) language spoken at home, and; (3) endorsement of gun safety to reduce gun violence. The model explained 18.2% of the variance in gun safety awareness (R² = 0.182, adjusted R² = 0.171). The final stepwise iteration did not result in a statistically significant improvement in model fit (ΔR² = −0.006, F change(1, 295) = 2.316, p = 0.129). Higher informal social control was significantly associated with higher gun safety awareness (β = 0.174, 95% CI [0.067, 0.281], aβ = 0.171, p = 0.001). Speaking Spanish vs. English in the home was also associated with higher awareness (β = 1.048, 95% CI [0.430, 1.666], aβ = 0.179, p < 0.001), as was endorsing gun safety to reduce gun violence (β = 0.189, 95% CI [0.121, 0.256], aβ = 0.314, p < 0.001; see Table 3).

## 5. Qualitative results

Two primary themes were identified through analysis: barriers to firearm safety engagement, and facilitators to firearm safety engagement. Theme 1, barriers to firearm safety engagement, included the following subthemes: (1) Insecure environment (structural; organizational), and; (2) Lack of meaningful engagement (organizational; cultural). Theme 2, facilitators to firearm safety engagement, included the following subthemes: (1) Location-specific contributors to safety (structural), and; (2) Collective identity and pride (communal). The intention of this analysis was to explore the perceptions of individuals rather than quantifying outcomes. The extent of quantifying results will be the use of the terms ‘most’ (more than half), ‘many’ (more than one-third), ‘some’ (less than one-third, more than 5), and ‘a few’ (between 3-5). Extracts from interviews are followed by the participant ID.

### 5.1 Theme 1: Barriers to firearm safety engagement

#### 5.1.1 Barrier subtheme 1: Insecure environment (structural; organizational)

Most participants reported infrastructure deficiencies that were perceived as signs of an insecure and dangerous environment that impeded their ability to engage with firearm safety, including a lack of street lights and sidewalks, and the frequency of hearing gunshots. These indicators of an insecure environment were reported as forcing Latino-Hispanic residents to focus on survival rather than firearm safety and feeling the need to protect themselves with guns. Participant #9 connected infrastructure deficiencies and poverty to potential gun use, stating, *“I live in an area where there’s a lot of poverty and no street lights. I wonder how much that plays a role [for] people feeling like they need to protect themselves with a gun.*”

Participants also described relations with law enforcement as contributing to a perceived insecure environment, describing the presence of police as uncomfortable and for some, even threatening. When discussing the role of police at a recent community event, participant #16 stated, *“Does it make me feel safer when we had [event] and we went to the park and saw the cops with all of their SWAT gear in the crazy show of force? That means more violence in the community.”* These participants reported that, in the wake of distrust and inactivity of police, Latino and Hispanic residents may decide to own or carry firearms for self-protection. When asked about firearm safety engagement among Latinos and Hispanics, participant #17 stated, *“A big part of [it] for any community is distrust with the police. What that means [is] this idea that we live in a different world, and people have bought into this notion that they need firearms to protect themselves.*”

#### 5.1.2 Barrier subtheme 2: Lack of meaningful engagement (organizational; cultural)

Current firearm safety initiatives were perceived by most as targeting audiences who already support prevention while missing harder-to-reach groups, including Latino-Hispanic gun owners and parents. Participant #2 reflected on a gun lock program at a recent community event, stating and expressed doubt that the program would reach parents who would benefit from it the most, stating, *“Honestly, I think sometimes that stuff falls on deaf ears. The people that stop to listen are probably going to be the people that would have already done it. I’m curious, how many people did they reach that it would actually pertain to?”* Many participants emphasized that a lack of culturally-specific firearm safety initiatives also impeded meaningful engagement, stressing the importance of acknowledging inequities which lead to Latinos and Hispanics being left out of firearm safety initiatives. When asked about the effectiveness of firearm safety in her community, participant #1 expressed concern over the absence of Latino-Hispanic nationality data, stating, *“I’d like to know specifically if the gun violence that has happened [was] carried out by Latinos. If so, were the people involved Latino or American? What is their nationality? Because they say it’s Latinos, but we don’t have a survey or anything that actually shows whether they are Latinos.”*

### 5.2 Theme 2: Theme 2: Facilitators of firearm safety engagement

#### 5.2.1 Facilitator subtheme 1: Location-specific contributors to safety (structural)

Many participants reported location-specific attributes that contributed to greater firearm safety, including a distinct rural identity and land stewardship. When asked what contributed to greater engagement with firearm safety in their community, participant #8 discussed the community’s history with a rural firearm culture which led to distinct firearm safety practices, stating, *“People who live in rural spaces are more likely to know how to use a gun safely. If you’re ranching, you probably own a shotgun, and you probably store it real safely. And you probably teach your kids young, ‘this is what a gun does. Don’t play with it.’”* The community was further distinguished by the presence of many longstanding multigenerational families, which was attributed to greater firearm safety engagement. When asked to share why he identified a specific area of the community as more likely to engage with firearm safety, participant #10 described some of the specific behaviors of multigenerational families that he perceived as keeping the community safe from firearm violence, stating, *“When you’re leaving to go to work, or when they’re coming back from somewhere, your family knows. They’re paying attention. They might say, ‘I noticed a different vehicle in your front yard’, or, ‘you left your garage door open.’ Having those kinds of connections make things safer.”*

#### 5.2.2 Facilitator subtheme 2: Collective identity and pride (communal)

Most participants reflected on aspects of their community’s shared identity that facilitated greater engagement with firearm safety, including a culture of knowing neighbors, interdependence, and pride. Participant #11 stated, *“Where I grew up, I knew all my neighbors and a good [amount] of the neighbors have been there since before I was born. Having that connection, and knowing who your neighbors are creates a better network of safety.”* For some participants, this sense of pride was connected to the visibility of Latino and Hispanic culture in the community, reporting cultural facets such as hearing Spanish spoken in public and the presence of multigenerational Latino-Hispanic families as signs of safety. Participant #11 described the connection between the community’s Latino-Hispanic culture and safety, stating, *“I think people tend to join together more in this community as far as Latinos. If you grew up here, and you’ve known somebody from [here], you know the culture, and it’s just safer because of the Latinos that lived here for generations.”*

A common thread across all interviews was participants’ reclamation of their community’s reputation through pride. These participants described the negative reputation of their community as a real and persistent part of their daily lives that they actively worked to challenge. When asked about firearm violence within their community, participant #18 stated, *“I never have experienced any violence in [community]… and again I worry, when I hear things like that, I worry that it’s a way to police the community, by using the excuse of gun violence.”* Most participants firmly rejected the stereotypes placed on Latinos and Hispanics in their community, instead emphasizing a self-defined collective identity which involved looking out for neighbors, working together to accomplish shared goals, and sharing pride. The act of reclaiming the community’s identity facilitated the development of community-specific firearm safety efforts and norms, and most participants expressed confidence that Latino and Hispanic residents could come together to address firearm violence. When asked about the community’s capacity to prevent gun violence, participant #7 pointed to several local initiatives, stating, *“There are amazing things that the community comes together for, like [community] pride day, and different events at the [location] and [event], where there is that sense of community. And so I think it’s possible to achieve more gun safety.”*

## 6. Mixed methods integrated results

The results from the quantitative and qualitative data are integrated using a joint display to inform the subsequent discussion (table 4). As illustrated, the left side provides quantitative results along with corresponding qualitative themes. The right side provides illustrative quotes, social ecological domains, and mixed methods inferences. First, the themes and subthemes relevant to firearm safety engagement derived from the qualitative portion of the study were mapped onto relevant findings from the survey portion of the study. Next, analyses were conducted to assess how well the quantitative findings mapped onto the qualitative themes and subthemes. This process was performed iteratively, and involved closely examining the quantitative findings, vetting the relevant qualitative themes, then returning to the quantitative data for support and confirmation for which qualitative concepts mapped well onto the quantitative items.

**Table 4.**
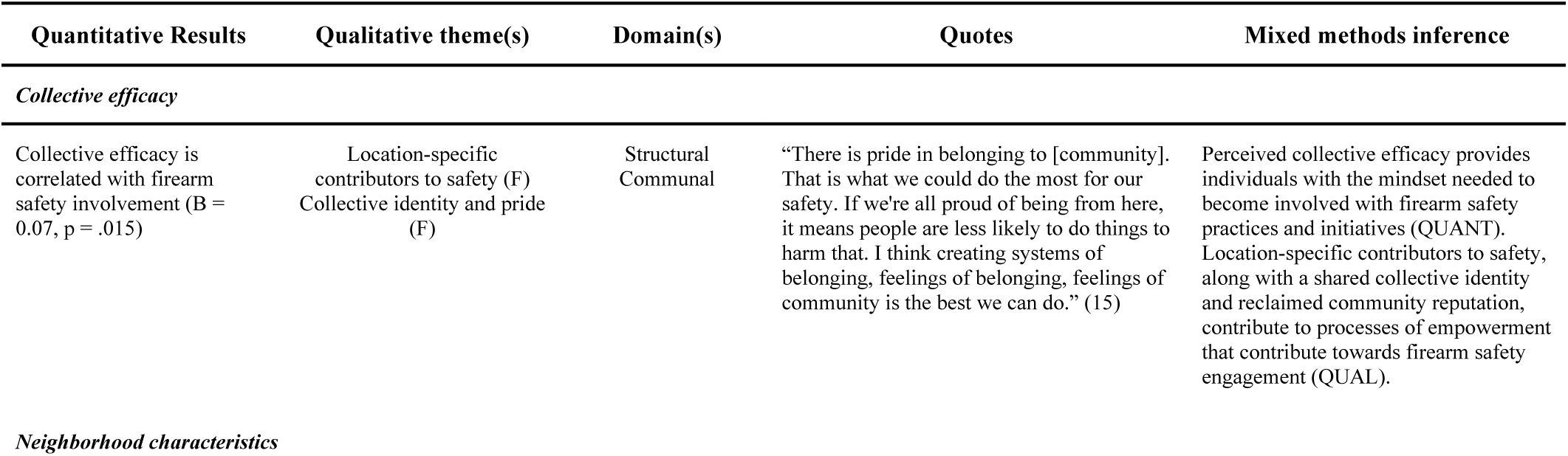
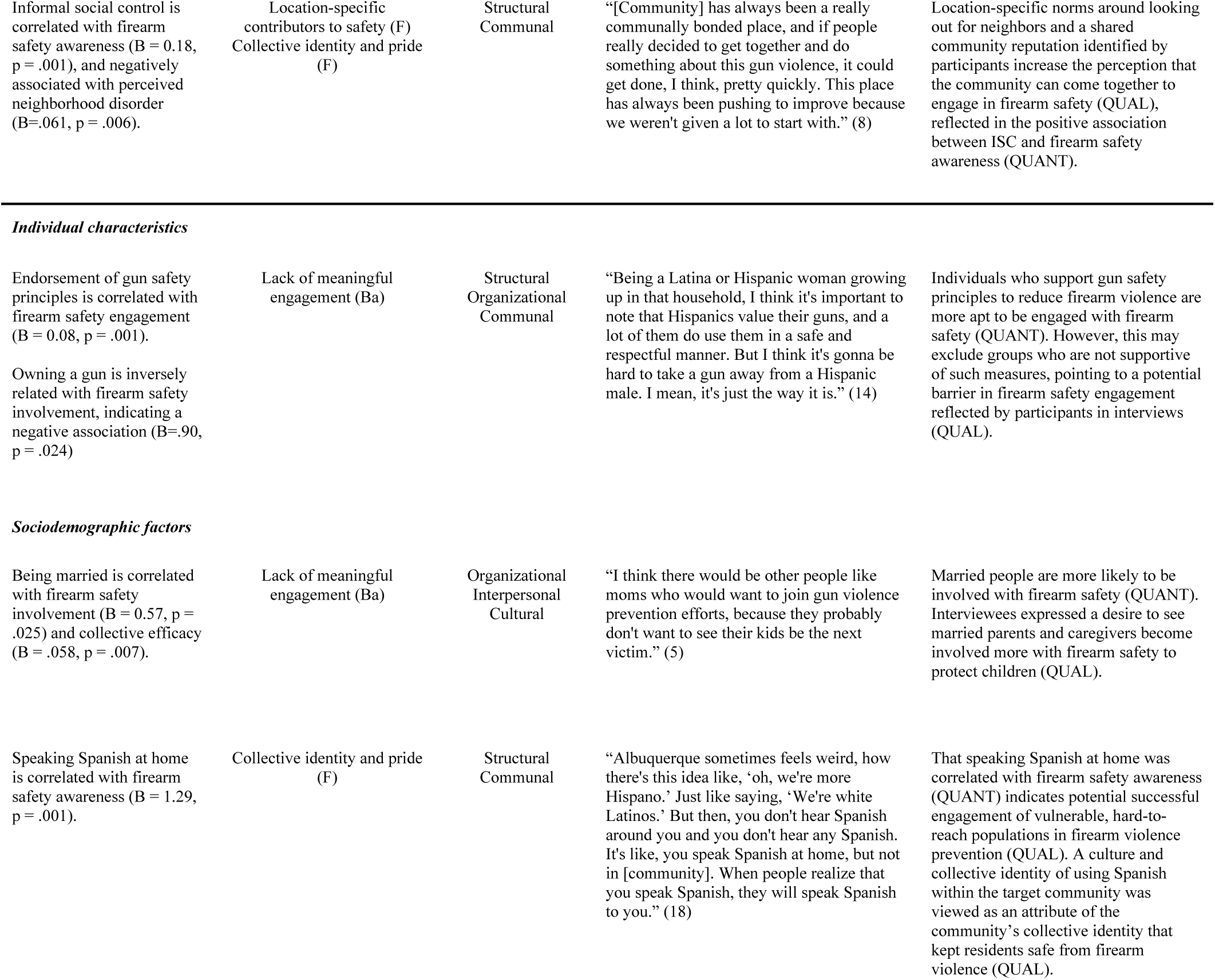
Joint Display of Mixed Methods Findings.

## 7. Discussion

This study explored the factors related to firearm safety engagement among a sample of Latino and Hispanic adults residing in New Mexico. Previous studies have stressed the importance of increasing firearm safety engagement within marginalized communities, but have seldom explored the potential factors affecting engagement. The findings expand prior research by considering the factors influencing firearm safety engagement among a marginalized and hard-to-reach population who have disproportionate experiences with firearm victimization, perpetration, and injury (94). The survey results indicate factors associated with participants’ individual firearm safety engagement, while the interview

data provides insights into the perceived barriers and facilitators across social ecological domains. The concurrent mixed methods study design helped acquire depth and breadth regarding the factors influencing firearm safety engagement among participants.

As hypothesized, Latino and Hispanic survey respondents who reported higher levels of collective efficacy and informal social control reported significantly higher firearm safety engagement. Qualitative findings expanded on these results by providing insights into the factors participants associated with collective efficacy. Interview participants expressed that location-specific attributes and a shared collective identity rooted in pride facilitated engagement with community-specific firearm safety practices. These practices included knowing and checking in on neighbors, rural firearm safety practices, and engaging with community-based initiatives to build cohesion and pride. Interviewees also articulated challenges with infrastructure and law enforcement that led to perceptions of living in an insecure environment, resulting in negative stereotyping of Latinos and Hispanics in their community and decreased firearm safety engagement.

By rejecting the negative reputation of their community while embracing a collective identity centered around pride, participants engaged in processes of empowerment to reclaim a shared identity different from what was assigned to them. Developing a shared collective identity and framing of social problems has been previously shown to empower groups to take prosocial actions (95). As this shared identity is embraced by increasing numbers of community members and connections are established, it could lead to exercises in collective empowerment, resulting in greater collective efficacy (96). This is evidenced by the majority of interview participants reporting that community-based initiatives and groups would be most effective in increasing firearm safety engagement and consequently, reducing firearm violence. Given these integrated findings, strategies for increasing firearm safety among Latinos and Hispanics should consider prioritizing place-specific and community-led initiatives aimed at enhancing collective efficacy by uplifting shared norms and values.

Survey results indicated that endorsing gun safety principles and not owning a gun were positively associated with firearm safety engagement, aligning with previous research indicating that firearm safety practices are more likely to be adopted by those who support gun safety principles (12). While it is commendable that firearm safety is being embraced by these groups, previous research has raised concern that such initiatives may not be reaching those at highest risk for experiencing firearm violence, including non-white gun owners (22). The findings of the present study echo this concern, as a significant negative correlation was found between owning a gun and firearm safety engagement. Interviews added depth to this finding, as participants reflected their concern that firearm safety initiatives would only reach demographics who are already supportive of them. The lack of engagement by groups deemed to be at higher risk for gun violence, including Latino-Hispanic gun owners, was viewed by most interviewees to be a barrier to firearm safety. In order to be successful among individuals with differing beliefs regarding gun ownership, future interventions should consider involving Latino-Hispanic gun owners in the formation and evaluation of potential firearm safety initiatives.

Among survey respondents, being married was correlated with greater firearm safety involvement, a finding that was only partially expanded upon by qualitative findings. The majority of interviewees reported a desire for parents and children to be more involved with firearm safety initiatives, expressing concern that current interventions were not reaching these groups. The common thread between these findings may be family cohesion, which has been previously found to exert a protective influence over unsafe firearm behaviors for Latinos and Hispanics (28). The desire to protect children from the known threat of firearm violence may create a rationale for Latino-Hispanic married people with children to engage with firearm safety as a measure of control. The findings of the present study indicate that marital status may be an aspect of family structure that influences differences in firearm-related behaviors, including greater engagement with safety practices. However, the survey used in this study did not ask questions regarding the presence of children in the home. To better understand this association and develop targeted interventions, future research should expand this finding by exploring the connection between marital status, family cohesion, and firearm safety engagement, examining distinctions based on race, ethnicity, and presence of children in the home.

Correlational and multivariate analyses indicated that speaking Spanish at home was associated with greater firearm safety engagement. This finding was extrapolated further in interviews, where some participants identified the frequent use of Spanish within their community as contributing to a greater sense of safety and pride. In one sense, these findings are contrary to previous literature, which have found a lack of Spanish language resources to be a barrier to health literacy regarding firearm safety (46). However, shared language use can be indicative of neighborhood racial homogeneity, which has been previously found to increase behaviors that benefit communities (31). For instance, a quantitative study using data from 3,868 household responses within 75 census tracts found that neighborhood racial homogeneity moderated and positively influenced the relationship between social cohesion and informal social control (31). Shared language use and neighborhood homogeneity may play a role in the present study due to the high percentage of Latinos and Hispanics residing in the target community.

While Spanish language use was associated with greater firearm safety involvement in the present study, qualitative findings indicate that a lack of meaningful engagement impedes the development of effective firearm safety initiatives. Many interviewees expressed concern that firearm safety initiatives were not being meaningfully engaged with by members of their community, expressing a desire for greater Latino-Hispanic input in the development of interventions. The results of the present study suggest that firearm safety interventions would be made more effective if they were developed with buy-in from Latinos and Hispanics to ensure that community-specific safety practices are prioritized. Future research should be conducted to elucidate the contribution of Spanish language and racial neighborhood homogeneity on firearm safety engagement among Latinos and Hispanics.

## 8. Limitations and Strengths

These findings reflect the experiences of Latino and Hispanic adults in one New Mexican community, limiting generalizability to other populations. The use of cross-sectional, non-random survey data introduces potential self-selection and social desirability bias, though anonymity and flexible completion options likely reduced these effects. Qualitative insights were limited by the small number of male participants and firearm owners, constraining subgroup comparisons. Despite these limitations, employing a mixed methods design strengthened the study’s rigor by enabling triangulation across data sources and expanding the depth and breadth of inquiry. The integrated approach offers a novel framework for understanding Latino-Hispanic engagement with firearm safety within complex social ecological contexts.

## 9. Conclusions

This timely and relevant mixed methods study written during a period of increasing firearm violence in the United States explored the social ecological factors related to firearm safety engagement among Latinos and Hispanics. Collective efficacy and shared communal identity were identified as potential facilitators for increasing firearm safety engagement, while perceptions of an insecure environment and lack of meaningful engagement were identified as potential barriers. Though extant literature has examined factors associated with firearm violence and gun ownership, firearm safety engagement among marginalized groups is an understudied area of inquiry. The mixed methods approach provided the opportunity to incorporate theoretical approaches fulsomely to the study design, methods, interpretation, and dissemination. This study’s findings signal the critical need for culturally specific and community informed intervention efforts to increase firearm safety engagement within Latino and Hispanic communities.

## Data Availability

To protect confidentiality of participants, data generated or analyzed during this study are limited to members of the research team and not available to the public.

## Acknowledgments

The author would like to thank the participants for trusting us with their stories about their communities and firearm violence prevention.

## Notes

### Competing Interest Statement

The authors have declared no competing interest.

### Funding Statement

This research received no specific grant from any funding agency in the public, commercial or not-for-profit sectors.

### Author Declarations

The research protocols were approved by the University of Toronto Human Research Ethics Unit. There was no patient involvement in the current study.

